# Quantitative Assessment of Aortic Hemodynamics for Varying Left Ventricular Assist Device Outflow Graft Angles and Flow Pulsation

**DOI:** 10.1101/2022.06.17.22276555

**Authors:** Akshita Sahni, Erin E. McIntyre, Jay D. Pal, Debanjan Mukherjee

## Abstract

Left ventricular assist devices (LVADs) comprise a primary treatment choice for advanced heart failure patients. Treatment with LVAD is commonly associated with complications like stroke and gastrointestinal (GI) bleeding, which adversely impacts treatment outcomes, and causes fatalities. The etiology and mechanisms of these complications can be linked to the fact that LVAD outflow jet leads to an altered state of hemodynamics in the aorta as compared to baseline flow driven by aortic jet during ventricular systole. Here, we present a framework for quantitative assessment of aortic hemodynamics in LVAD flows realistic human vasculature, with a focus on quantifying the differences between flow driven by LVAD jet and the physiological aortic jet when no LVAD is present. We model hemodynamics in the aortic arch proximal to the LVAD outflow graft, as well as in the abdominal aorta away from the LVAD region. We characterize hemodynamics using quantitative descriptors of flow velocity, stasis, helicity, vorticity and mixing, and wall shear stress. These are used on a set of 27 LVAD scenarios obtained by parametrically varying LVAD outflow graft anastomosis angles, and LVAD flow pulse modulation. Computed descriptors for each of these scenarios are compared against the baseline flow, and a detailed quantitative characterization of the altered state of hemodynamics due to LVAD operation (when compared to baseline aortic flow) is compiled. These are interpreted using a conceptual model for LVAD flow that distinguishes between flow originating from the LVAD outflow jet (and its impingement on the aorta wall), and flow originating from aortic jet during aortic valve opening in normal physiological state.

## 1 Introduction

Heart failure continues to be a global public health concern, affecting at least 26 million individuals globally, and being recognized as an emerging epidemic as of 1997 ^[9,47,46]^. Within the United States alone, over 6 million individuals above the age of 20 have advanced heart failure ^[8]^, leading to an estimated economic burden of over 30 billion USD on the US healthcare system ^[12]^. Left ventricular assist devices (LVADs) are mechanical pumps that offer circulation support in patients with advanced heart failure by shunting blood from dysfunctional left ventricle directly into the aorta. Through recent design advancements, LVADs have emerged as a primary treatment modality for heart failure as both bridge-to-transplant as well as destination therapy in patients not suitable for a transplant^[51]^. Despite advancements in LVAD therapy, patient outcomes on LVAD remain associated with significant morbidity and mortality. Stroke and gastro-intestinal (GI) bleeding are the leading complications post LVAD therapy. Reported stroke rates after LVAD implantation are anywhere between 11-47 percent ^[37,21,42,1]^, while incidence of GI bleeding in patients on LVAD is reported to be around 11-40 percent ^[25,34,48]^, going as high as 60 percent in some cases ^[31]^. Factors that influence post-implant stroke and bleeding in LVAD patients are not well understood pre-implant. It is widely regarded that the altered state of hemodynamics post-LVAD implantation, as compared to the baseline physiological flow pre-implantation, plays a crucial role in determining risk and etiology of complications in patients. Accurate quantification of spatiotemporally varying hemodynamic features can, therefore, be critical to assess treatment efficacy ^[6,50]^.

Computational flow modeling has emerged as a suitable avenue for comprehensive characterization of hemodynamics post-LVAD implantation ^[16]^, as documented in a range of studies. The effects of the LVAD out-flow graft surgical attachment angle on hemodynamics in the aortic arch and thromboembolic potential has been extensively studied ^[32,23,28,10,4,43]^. Several specific aspects have also been investigated computationally, such as: the role of pulse modulation of LVAD flow studied across adult and pediatric population ^[55,56,13]^; additional VAD surgical attachment parameters ^[11]^; the role of viscoelastic hemorheology in VAD-driven circulation ^[20]^; and the effect of intermittent aortic valve reopening ^[29]^. However, few key hemodynamic aspects remain incompletely understood, which motivate our study presented here. First, quantitative characterization of the extent of hemodynamic alteration with reference to a *baseline flow* pre-implantation needs attention. Second, while majority focus has been on flow in the aortic arch region due to their influence in governing stroke risks, it is important to characterize the altered state of hemodynamics at locations distal from the outflow graft. This is especially important for the abdominal aorta region, owing to the potential underlying hemodynamic role in GI bleeding risks. Third, there is a need to identify a fundamental flow physics interpretation, that can enable a unified generalizable understanding of the altered state of hemodynamics, beyond individual patient-specific spatiotemporally varying flow trends. To address these goals, we present a systematic *in silico* hemodynamic study for varying LVAD graft surgical parameters, and flow modulation; and compare the LVAD-driven flow to an estimate of the baseline flow without an LVAD, through a series of commonly used hemodynamic descriptors to characterize extent of hemodynamic alteration when compared to the baseline flow.

## 2 Methods

### 2.1 Image-based modeling

A patient-specific vascular network comprising the aortic arch and branch arteries extending up to the iliofemoral arteries was obtained from computed tomography (CT) images (for model details refer to Sim-Vascular ^[53] [49]^ and associated vascular model repository ^[45]^, as well as our prior work ^[38]^). The CT images were segmented using SimVascular’s built-in 2D lofted image segmentation technique, and converted into a 3D triangulated surface representing the artery lumen for the *baseline model* without an LVAD. Subsequently, an image-based geometric modeling procedure was devised for virtual surgical placement of an LVAD outflow graft, as outlined in Figure 1, panel a. First, an additional pathline for guiding LVAD outflow graft placement was generated, along which a series of circular segmentations of diameter equal to the graft were created. These segmentations were lofted to generate a surface model for the graft. The surface was then iteratively adjusted by translating and deforming the paths and segments to ensure the resulting graft does not intersect nearby organ (heart, lungs) or bone (sternum, ribs) boundaries - thereby generating realistically achievable surgical LVAD outflow graft anastomoses. The angle between the final pathline for the graft, and the pathline for aorta, were noted in terms of: (a) orientation towards or away from the aortic valve (labelled with an *Inc* tag); and (b) orientation towards left or right of the heart across the coronal plane (labelled with an *Azi* tag). For this study, we considered a set of 9 different parametric graft anastomoses, based on 3 combinations of the 2 angles each. For the former, we chose graft anastomosis angles: (1) perpendicular to the aorta (*Inc90*); (2) 45° towards the aortic valve (*Inc45*); and (3) 45° towards the aortic arch (*Inc135*). For the latter, we chose graft anastomosis angles: (1) 45 ° right of the heart (*AziNeg45*); (2) perpendicular to the coronal plane (*Azi0*); and (3) 45 ° left of the heart (*Azi45*). These resulting 9 combination anastomoses are illustrated in Figure 1, panel b., leading to a set of 10 vascular models including the *baseline model* of the aorta without an operational LVAD.

**Figure 1:**
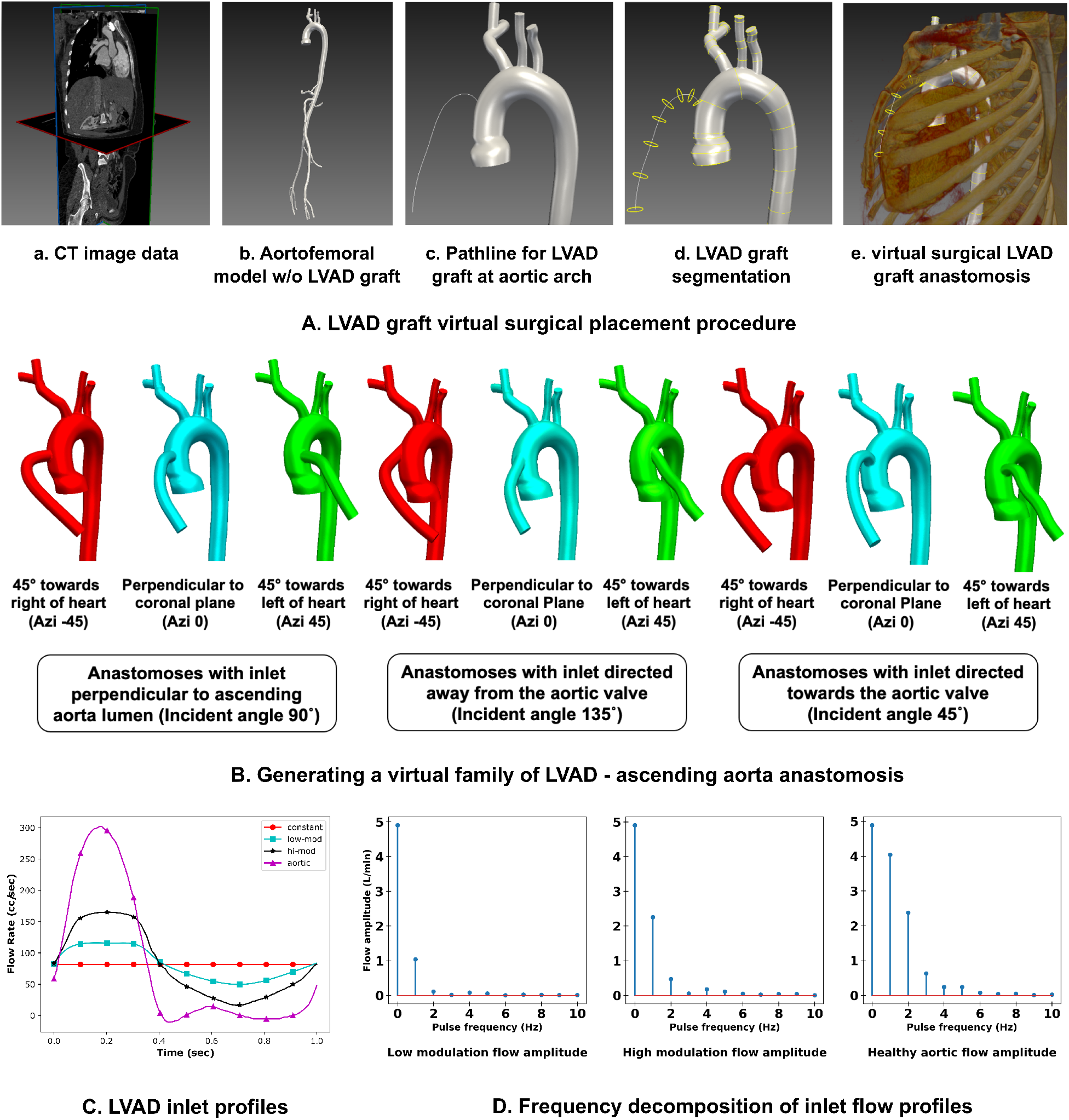
Illustration of modeling methodology details. Panel a. demonstrates the workflow for image-guided graft placement, using SimVascular image-processing toolkit. Panel b. depicts the 9 different graft anastomoses, created by varying graft attachment angles towards/away from the heart, and towards/away from the aortic valve (see Section 2.1). Panel c. illustrates the various LVAD flow profiles used for this study, including a frequency domain decomposition of pulse-modulated signals compared to baseline.

### 2.2 Computational hemodynamics modeling

Blood was assumed to be a Newtonian fluid with an effective density of 1,060 kg.m^3^ and effective viscosity 0.004 Pa.s. The computational domain for each of the 10 models outlined in Section 2.1 was discretized into a mesh comprising linear, tetrahedral elements with a maximum edge size of ≈0.67 mm. Assuming rigid vessel walls, 3-dimensional blood flow through all models was computed using the Navier-Stokes equations and continuity equation for blood momentum and mass balance respectively. These equations were solved using a Petrov-Galerkin stabilized linear finite element method (in the SimVascular suite) ^[53]^ to compute space-time varying velocity and pressure fields in the arterial pathway. Flow in the baseline model was driven by assigning a physiologically measured pulsatile flow profile at the aortic root inlet, mapped spatially into a Womersely-type flow profile across the inlet face. For the 9 models with LVAD graft, the aortic root inlet was assumed to be closed and modeled as a rigid wall. This assumption is based on existing studies that indicate that during periods of high LVAD support the aortic valve remains continuously closed ^[27,30,52,15]^; and any trans-valvular flow is only intermittent, contributing a lower level of flow ^[29]^. Flow was instead driven by assigning an inflow profile to the LVAD outflow graft entrance, mapped spatially into a Womersley-type flow profile across the inlet. Three different scenarios of LVAD flow were considered: (a) a constant uniform flow over time; (b) flow with a low extent of pulse modulation; and (c) flow with a high extent of pulse modulation; with the pulse modulated flow profiles adapted from prior studies ^[24]^. The time-averaged inlet flow was fixed at 4.9 liters/min for all scenarios. Figure 1, panel c. and Table 1 together illustrate the three profiles, their frequency component contributions, and their pulsatility index (PI) defined as:

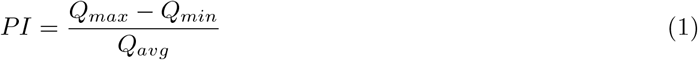

**Table 1:** A comparison of the pulse-modulated LVAD flow profiles with that of baseline aortic inflow profile used in this study.

| Pulse profile | Mean flow (l/min) | Range (l/min) | Pulsatility Index (PI) |
| --- | --- | --- | --- |
| low modulation | 4.90 | 3.0 - 7.0 | 0.81 |
| high modulation | 4.90 | 1.0 - 10.0 | 1.82 |
| aortic | 4.90 | -0.6 - 18.0 | 3.83 |

with *Q*_*max*_ being the maximum flow rate; *Q*_*min*_ being the minimum flow rate; and *Q*_*avg*_ the average flow rate for the LVAD. For the 9 LVAD anastomoses models, each of the 3 different LVAD flow pulsation cases were simulated, leading to a total of 28 parametric hemodynamics simulations (including the baseline model). For each simulation, the numerical integration over time was conducted using a time-step of 0.001 sec. The simulations were run for three pulse cycles allowing for sufficient flow development in time (as demonstrated in prior works ^[39]^). Effects of the truncated downstream vasculature on the flow was incorporated using 3-element Windkessel boundary conditions at each outlet of the computational domain. Individual Windkesel parameters: proximal resistance (*R*_*p*_); distal resistance (*R*_*d*_); and compliance (*C*); were computed based on assumed target flow distribution across each outlet obtained from existing literature ^[45]^. The Windkessel parameters were kept the same across all 10 models, and the values used here have been listed in Table 2.

**Table 2:** A complete list of all boundary condition parameter values for three-element Windkessel boundary conditions assigned at all model branch outlets.

| Artery name | $R_p$<br>[gm.cm<sup>-4</sup>.s<sup>-1</sup>] | $C$<br>[gm<sup>-1</sup>.cm<sup>4</sup>.s<sup>2</sup>] | $R_d$<br>[gm.cm<sup>-4</sup>.s<sup>-1</sup>] |
| --- | --- | --- | --- |
| Right Subclavian | 788.82 | 0.0002090 | 13297.26 |
| Right Common Carotid | 6697.69 | 0.0001235 | 17222.63 |
| Left Common Carotid | 6708.98 | 0.0001235 | 17251.66 |
| Left Subclavian | 788.87 | 0.0002090 | 13298.17 |
| Hepatic | 1114.61 | 0.0001456 | 18789.07 |
| Splenic | 1086.33 | 0.0001456 | 18312.39 |
| Superior Mesenteric | 829.30 | 0.0001970 | 13979.66 |
| Left Renal | 4102.96 | 0.0001970 | 10550.48 |
| Right Renal | 4141.67 | 0.0001970 | 10650.01 |
| Left Internal | 3108.73 | 0.0000529 | 52404.23 |
| Right Internal | 3109.10 | 0.0000529 | 52410.58 |
| Left Profunda | 6620.49 | 0.0000247 | 111602.55 |
| Right Profunda | 6570.55 | 0.0000247 | 110760.65 |
| Left External Circumflex | 3316.24 | 0.0000494 | 55902.32 |
| Right External Circumflex | 3299.25 | 0.0000494 | 55615.95 |
| Left Femoral | 3322.05 | 0.0000494 | 56000.20 |
| Right Femoral | 3312.96 | 0.0000494 | 55847.04 |

### 2.3 Quantification of hemodynamic descriptors

Quantitative analysis of aortic hemodynamics across the 28 parametric simulations was conducted by post-processing the flow velocity data over the third simulated cardiac cycle. Customized workflows were created for post-processing flow data using the open-source visualization application Paraview, and the underlying Visualization Toolkit (VTK) library ^[2]^. A set of three Eulerian flow descriptors were devised based on quantities widely utilized in studying arterial hemodynamics. These descriptors were computed for: (a) the aortic arch until the descending aorta; and (b) the abdominal aorta beyond the descending aorta until the iliofemoral branch. The first descriptor focused on the state of helical swirling flow in the aorta. Curvature and tortuosity of the aorta causes development of helical flow patterns physiologically observed in healthy aortic hemodynamics ^[3,19]^. Helical flow is acknowledged to be physiologically beneficial with respect to normal organ perfusion, equalizing aortic wall shear stress distribution, and reducing flow stagnation ^[17,36,18,14]^. Helical flow is commonly described using the flow helicity, defined based on flow velocity ***<u>u</u>*** as follows:

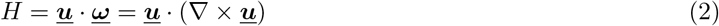

Here, flow helicity is further re-interpreted in terms of a scaled indicator of the local alignment between flow velocity and vorticity, commonly defined as the Local Normalized Helicity (LNH) as follows:

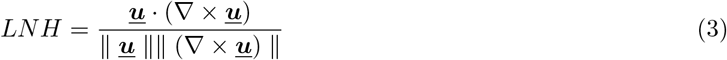

LNH has been widely used in visualization of complex flow patterns in cardiovascular flows ^[35,18,19]^. Here, we used a time averaged Local Normalized Helicity (LNH) across one cardiac cycle, defined as below, as a helical flow descriptor compared across all of the parametrically varied simulation cases:

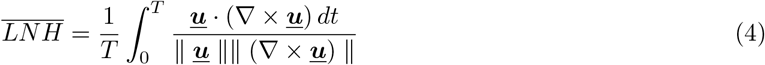

Based on 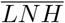 definition in Equation 4 we further devised a volume integrated helicity metric Φ_*H*_ and volume averaged helicity metric ⟨Φ_*H*_⟨ defined as follows for a given range of averaged 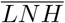 values [*L*_1_, *L*_2_]:

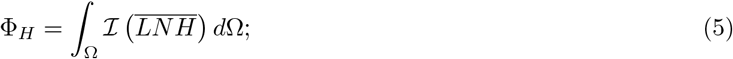

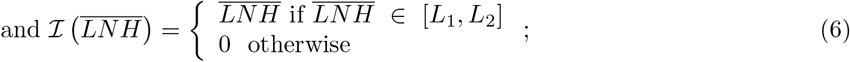

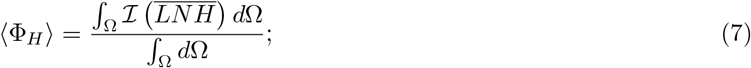

where integration domain Ω denotes the aortic arch region or abdominal aorta region as stated earlier. The second descriptor for our analysis focused on the extent of vortical structures formed in the aorta due to jet flow emanating from the LVAD graft, as reported in previous studies undertaking qualitative assessment of LVAD induced aortic flows ^[57]^. Here, a quantitative description of spatiotemporally varying vortex structures is conducted using the widely adopted *Q*-criteria, which identifies locations where rotational flow dominates straining flow ^[26]^ as outlined below:

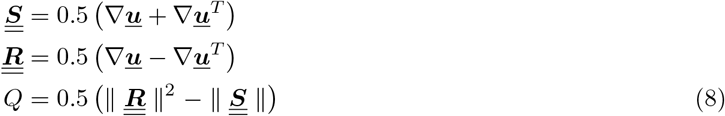

where the tensor 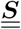 denotes state of strain in the flow, 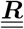 denotes the state of rotation in the flow, and the final derived *Q*-criteria in Equation 8 denotes a vortex to be the spatial region in the flow where *Q* > 0. For more specific quantitative comparison across all the cases studied here, we devised a volume integrated Q-criterion metric Φ_*Q*_ and volume averaged Q-criterion metric ⟨Φ_*Q*_⟩ defined as follows:

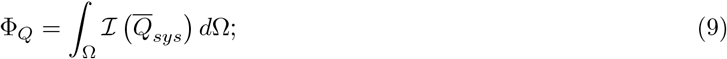

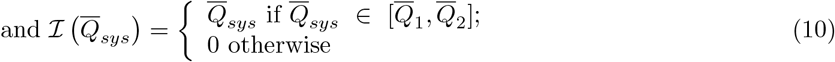

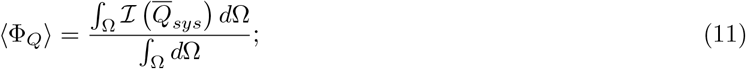

where Ω denotes the integration domain, which is either the aortic arch region or the abdominal aorta region (as stated earlier); 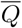 denotes the *Q*-value normalized against the systolic *Q*-value for the baseline model; ℐ denotes an indicator function to isolate the 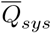 value between the range of 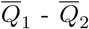. The third descriptor focused on the state of wall shear stress (WSS), a widely studied hemodynamic descriptor used to characterize the effect of flow disturbances on the mechanobiology of the endothelial cell layer ^[5]^. High WSS can cause significant decline in the endothelial function and may evoke phenotypic changes linked to thrombosis, hemostasis and inflammation ^[22,54]^. Both lower and higher (compared to physiological levels) wall shear stresses have been linked to atherosclerotic plaque development ^[33]^. Wall shear stress is computed based on flow velocities proximal to the wall with a normal ***n*** using the relation:

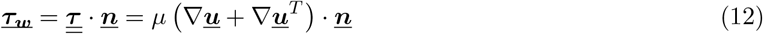

where *τ*_*w*_ is the WSS, and *μ* is dynamic viscosity of the fluid. Here, the time averaged wall shear stress (TAWSS) magnitude was compared across the parametrically varied simulation cases, estimated using the following relation:

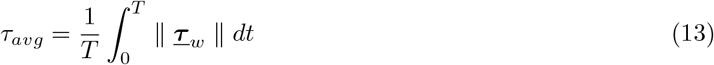

where for time averaging, we use *T* = one pulse cycle. For the aortic arch the TAWSS is compared for the arch surface excluding the LVAD graft and branching vessels; and similarly for the abdominal aorta, the TAWSS is compared for the aorta surface excluding all branching vessels - for all flow scenarios considered. A surface-averaged TAWSS descriptor was also computed, defined as follows:

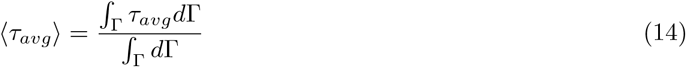

where Γ denotes the surface of the aortic arch and the abdominal aorta, without any of the branching arteries or the LVAD graft (for the arch cases). Furthermore, comparison between TAWSS in LVAD cases vs. baseline flow was conducted by calculating a TAWSS difference as follows: (a) WSS contribution from the LVAD graft and the first generation branches (carotid and subclavian arteries) was removed; (b) for the LVAD cases, the aortic arch WSS values were resampled to match the WSS samples in the baseline aortic arch; (c) WSS for each LVAD case was projected onto the baseline wall to calculate the TAWSS difference computed on the surface, as shown in the equation:

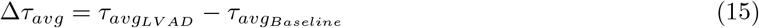

Lastly, the above TAWSS estimates were used to conduct a quantitative comparison of TAWSS difference by taking a surface average over the aortic arch surface as follows:

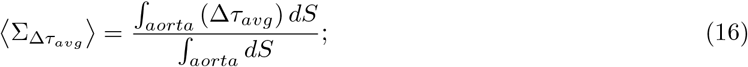

## 3 Results

### 3.1 Helical hemodynamics driven by the LVAD

Simulated blood flow patterns for varying LVAD graft anastomoses and varying LVAD pulsation have been visualized using streamtubes illustrated at peak systole in Figure 3. Panel a. corresponds to the aortic arch, and panel b. corresponds to the abdominal aorta region. The systole was at 0.18 seconds (that is, instance of peak LVAD flow in pulse modulated cases). Each row in panels a. and b. denotes graft orientation towards/away from aortic valve (*Inc* angles); and columns denote pulse modulation categorized per graft orientation across the coronal plane (*Azi* angles) as detailed in Section 2.1. Corresponding baseline flow is illustrated in Figure 2, where panel a. depicts the systolic velocity streamtubes. Several key aspects of the flow are observed from Figure 3. First, impingement of the LVAD outflow jet onto the aorta wall drives the aortic hemodynamics in presence of a non-functional aortic valve. Based on LVAD geometry, fluid parameters, and imposed mean flow-rates as in Section 2.2, the average flow velocity based Reynolds number on the aorta was 1400, with the highest Reynolds numbers being up to 3000 in the jet impingement region. Second, helical aortic flow patterns are present for the baseline as well as each of the 27 LVAD flow scenarios simulated. Third, for increasing LVAD outflow graft angles away from the valve (*Inc* angles), a prominent flow stasis region is observed at the root of the proximal aortic arch as the impingement site advances away from the root. Together, these illustrations qualitatively identify differences in aortic hemodynamics compared to the baseline flow, with varying LVAD outflow graft angles as well as pulsation. The extent of helical flow, and trends across varying graft anastomoses and pulse modulation, is further characterized using the 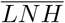 values computed as described in Section 2.3, illustrated in Figures 4 and 5 for aortic arch and abdominal aorta respectively. Panel a. in Figures 4 and 5 illustrate helical flow for the 27 LVAD flow scenarios using 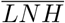 isosurfaces taken at 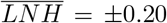, which corresponds to an averaged angle between velocity and vorticity vectors of ≈78° (positive) and ≈168° (negative) respectively. Row and column arrangement with respect to angles and pulsation is the same as Figure 3. Comparing these with 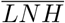 for the baseline model (shown in Figure 2, panel b.), we qualitatively observe that LVAD flow alters the extent of helical flow in the arch and abdominal aorta (compared to normal aortic flow). Furthermore, we observe that helical flow in the aortic arch is strongly influenced by both of the LVAD graft anastomoses angles as well as pulse modulation. These inferences are further established quantitatively by comparing the volume integrated helicity metric Φ_*H*_ (as outlined in Section 2.1), indicating the extent of positive or negative helical flow for the 27 LVAD flow scenarios, for both aortic arch and abdominal aorta regions. For the arch, the integral in Φ_*H*_ is computed across the arch excluding, contributions from the LVAD outflow graft, and branching vessels. Likewise, for the abdominal aorta, this is computed across the aorta excluding contributions from the branching vessels. Panel b. in Figures 4 and 5 presents the ratio of ⟨Φ_*H*_⟩ for all 27 LVAD cases against that for the baseline case for [*L*_1_, *L*_2_] = [0.2, 1.0]; and panel c. depicts the same ratio for [*L*_1_, *L*_2_] = [−1.0, −0.2]. Together, these illustrations present the variation of ⟨Φ_*H*_⟩ with graft angles and pulsation, when compared against baseline aortic flow without LVAD. We observe that while the baseline flow is more negatively helical, flow through the LVAD graft leads to aortic hemodynamics being more positively helical for both the arch and the abdominal aorta (compared to the baseline); with the extent of positive helicity stronger in the abdominal aorta than the arch. For positive helical flow, the effect of graft angles was less pronounced than pulsation. Specifically, for all cases, pulsed LVAD scenarios led to lesser extent of positive helicity, than constant LVAD flow; an effect that is more pronounced for flow at the abdominal aorta than the aortic arch. For negative helicity, the variation with pulsation was less pronounced, but variation with graft angle was more consistent. Specifically, for the aortic arch, the extent of negative helicity reduced across *Azi* angles of 0°, 45°, and −45°; while for the abdominal aorta this trend is generally reversed. At the aortic arch, for all models considered, most negative helicity occurs in *Inc135Azi0* anastomosis, and most positive helicity in *Inc135Azi-45* anastomosis, both constant LVAD flows. Likewise, for the abdominal aorta, most negative helicity occurs in *Inc90Azi-45* anastomosis, while most positive helicity occurs in *Inc90Azi45* - again both being constant LVAD flows.

**Figure 2:**
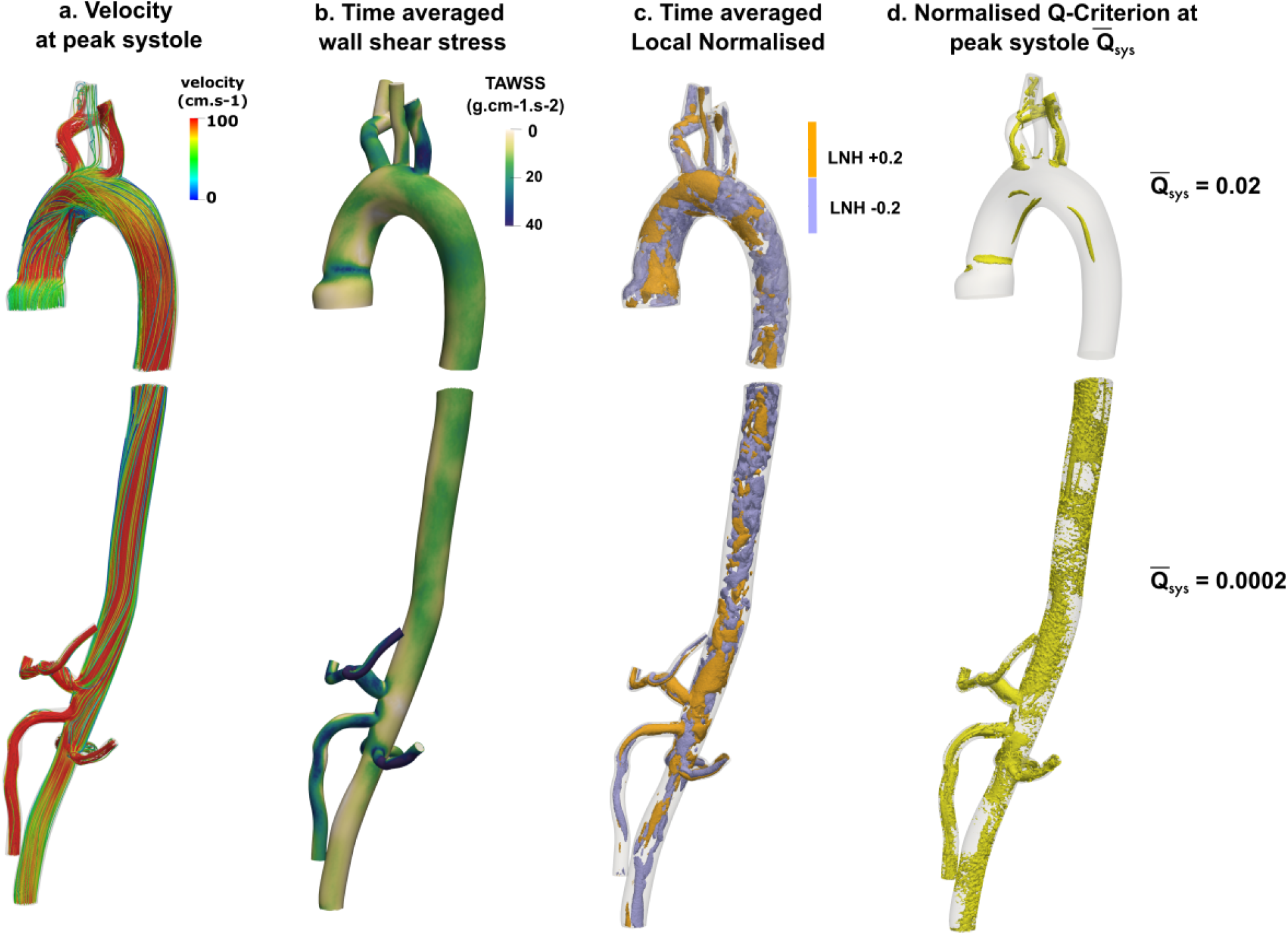
Computed hemodynamics for baseline model (that is, without LVAD). Panels a.-d. depict velocity streamtubes viewed at peak systole (see also Figure 1,panel c.); time averaged wall shear stress; iso-surfaces of time averaged LNH viewed at ±0.2; and normalised Q-criterion. The aortic arch region, and abdominal aorta region are shown separately.

**Figure 3:**
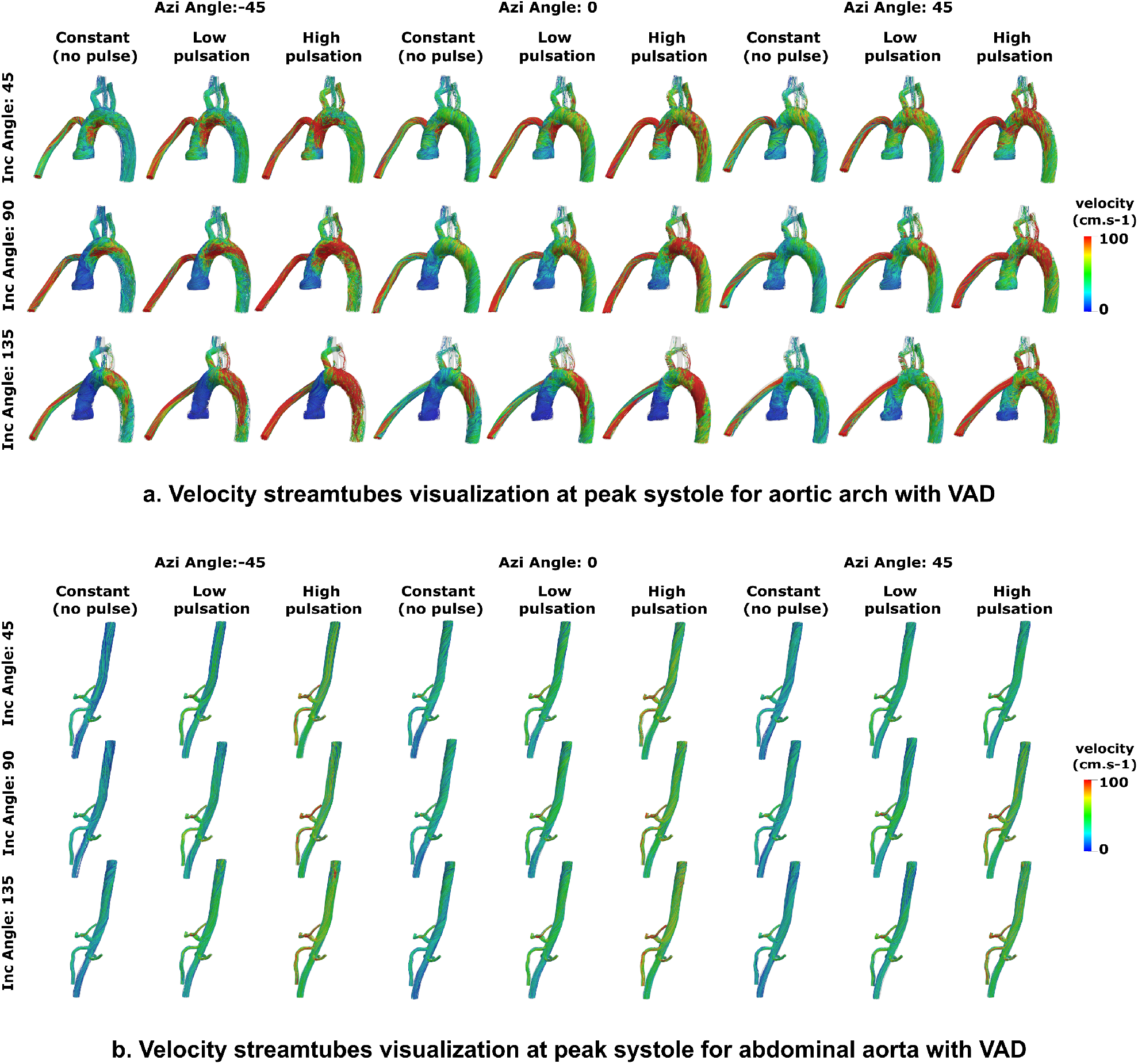
Illustration of blood flow velocity at t=0.18 seconds in a 1 second cycle window (for pulse modulated cases), for all 27 LVAD flow scenarios considered. Panel a. depicts the streamtubes for the aortic arch region, panel b. depicts the same for the abdominal aorta region.

**Figure 4:**
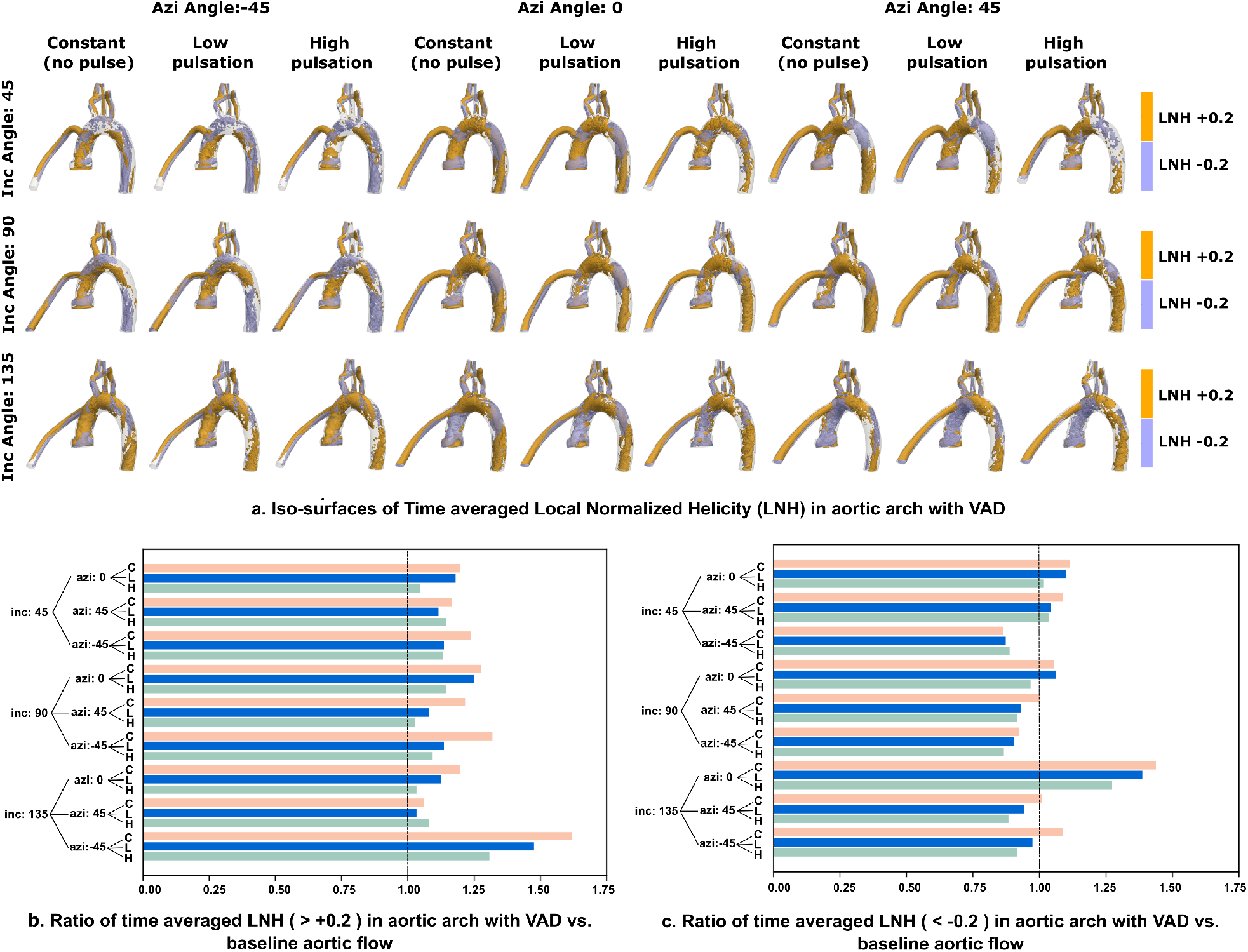
Simulated helical flow descriptors for the aortic arch region. Panel a. presents the time averaged LNH iso-surfaces at ±0.2. Panels b. and c. present the ratio of the computed helicity descriptor ⟨ Φ_H_ ⟩ for each of the 27 LVAD scenarios as compared against baseline flow. Ratio=1.0 (marked on panels b.,c.) represents baseline flow without LVAD.

**Figure 5:**
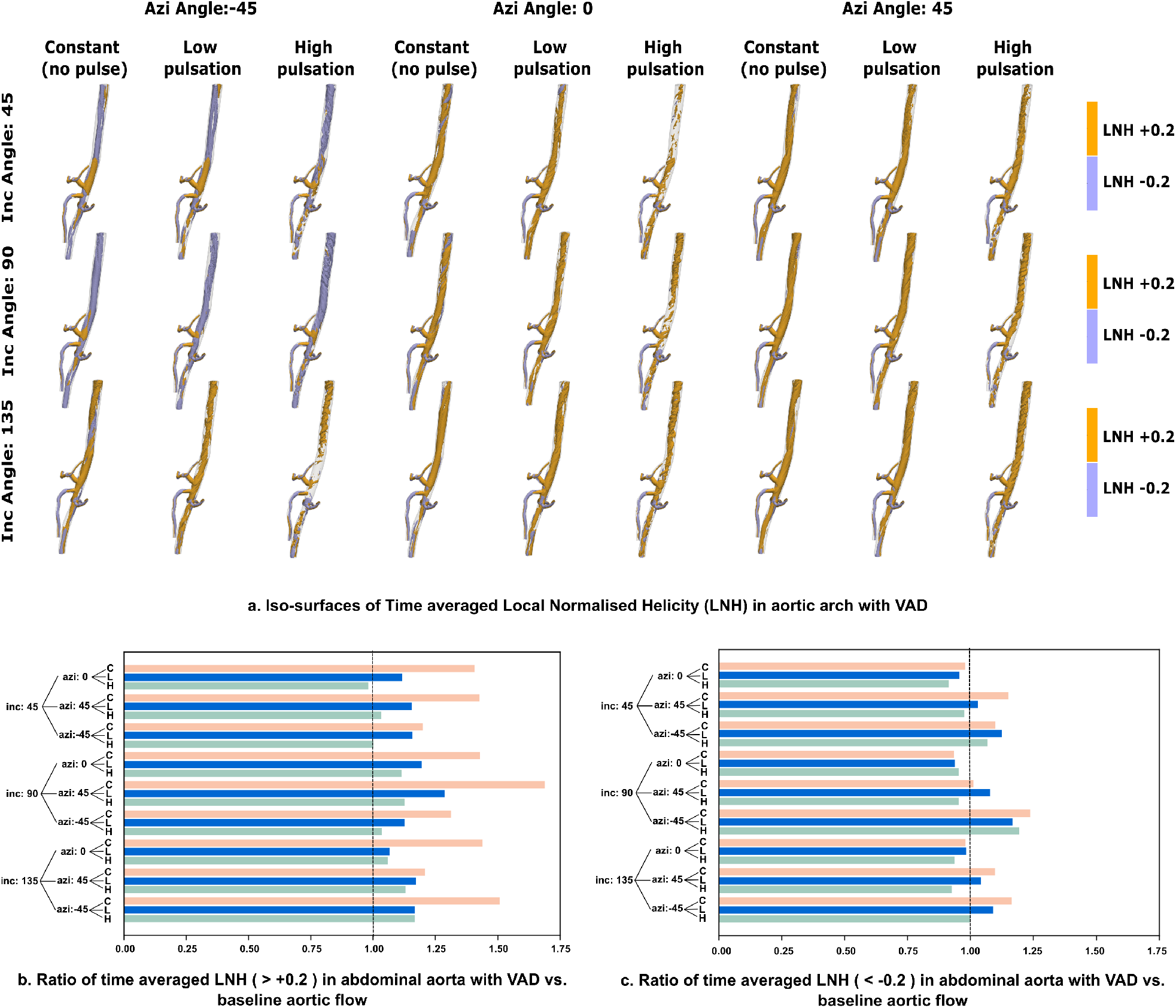
Simulated helical flow descriptors for the abdominal arch region. Panel a. presents the time averaged LNH iso-surfaces at ±0.2. Panels b. and c. present the ratio of the computed helicity descriptor ⟨Φ_H_⟩ for each of the 27 LVAD scenarios as compared against baseline flow. Ratio=1.0 (marked on panels b.,c.) represents baseline flow without LVAD.

### 3.2 Vortex structures in LVAD hemodynamics

Figure 6, panels a. and c. illustrate the extent of vortical structures for all 27 LVAD flow scenarios considered using *Q*-criterion isosurfaces (see Section 2.3), presented for the aortic arch and abdominal aorta respectively. Row and column arrangement with respect to angles and pulsation is the same as Figure 3. For the aortic arch, iso-surfaces were taken at 2 percent of the maximum *Q*-criterion value for the baseline case (evaluated to be 2.9 × 10^6^ at peak systole), and for the abdominal aorta, iso-surfaces were set at a much lower value of

**Figure 6:**
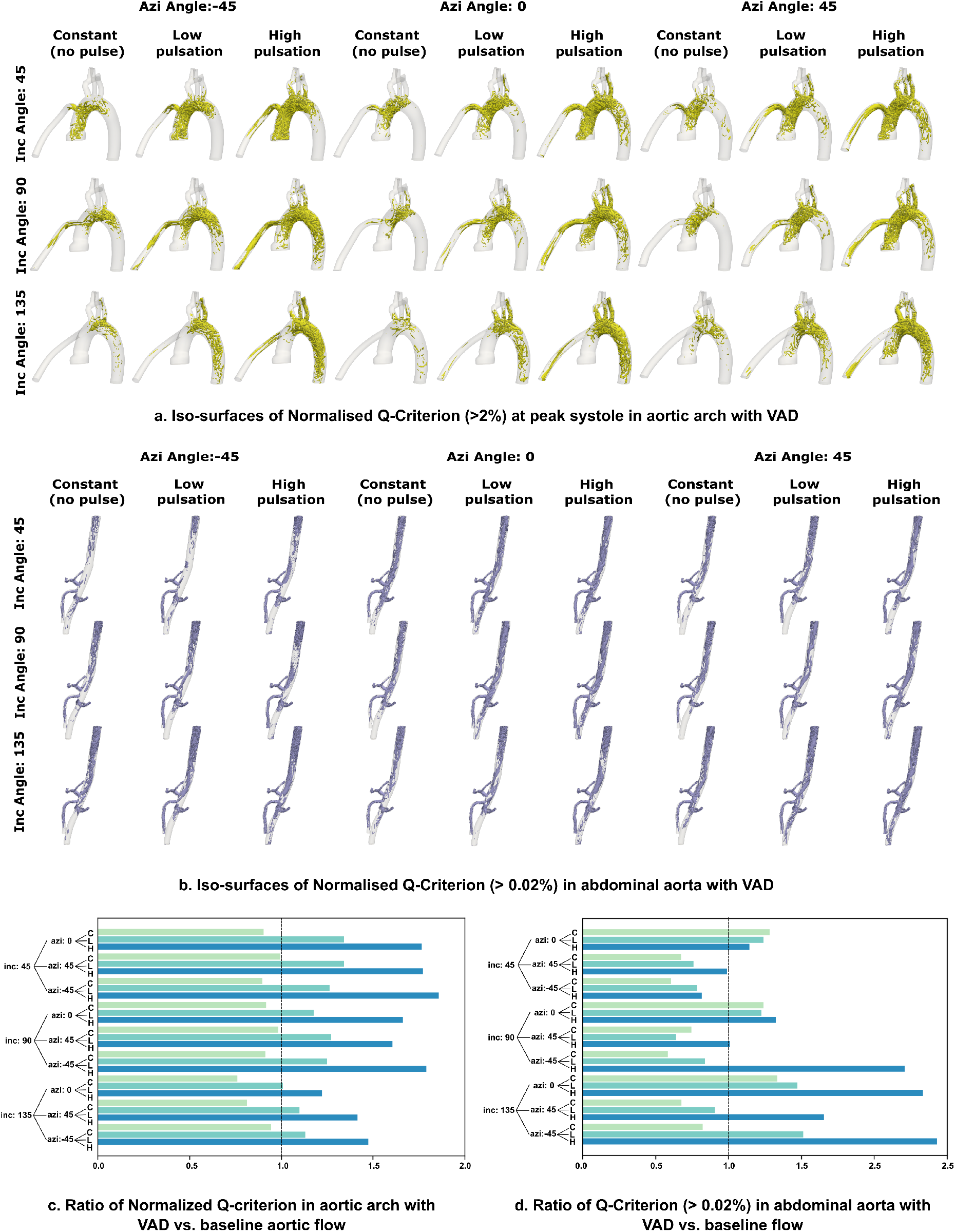
Illustration of vortex structures and extent of vorticity in all 27 simulated LVAD flows. Panel a. presents iso-surfaces of normalized Q-criterion taken at 0.02 for the aortic arch, while panel c. presents the same for the abdominal aorta with iso-surfaces taken at 0.0002. Panels b. and d. illustrate the ratio of computed vorticity descriptor ⟨Φ_Q_⟩ for all 27 LVAD flow scenarios, compared against baseline flow. Ratio=1.0 (marked on panels b.,d.) represents baseline flow without LVAD.

0.02 percent. Corresponding baseline systolic values are presented in Figure 2, panel d. When compared to the baseline case, the extent of vorticity in the aortic arch due to the LVAD flow is significantly higher as the outflow jet from the LVAD graft impinges on the aorta wall. With increasing pulsation, the extent of vortical structures increases, spanning into a greater region of the vasculature towards the descending thoracic aorta into the abdominal aorta. Additionally, as the anastomosis angles away from the aortic valve, the presence of a stagnation flow at the aortic root is evidenced by the lack of any vortical structures as seen in Figure 6 panel a. This is consistent with our observations of velocity streamtube patterns in Section 3.1. These inferences are further advanced quantitatively by comparing the volume integrated and averaged Q-criterion metric ⟨Φ_*Q*_⟩ (as defined in Section 2.3) for varying anastomosis angles and pulsation, for both the arch and abdominal aorta. For the aortic arch, ⟨Φ_*Q*_⟩ was computed for the arch excluding the LVAD outflow graft and the first generation branching arteries. 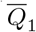 was set at 0.02, and 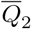 was set to the maximum 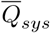 ratio for the model. For the abdominal aorta, ⟨Φ_*Q*_⟩ was computed excluding all the first generation branches from the aorta. 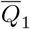 was set to a much lower value of 0.0002, while 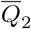 was set to the maximum ratio. Figure 6 panels b. and d. depict the ratio of ⟨Φ_*Q*_⟩ in the arch and abdominal aorta respectively, for flow with an LVAD vs. baseline aortic flow, for all 27 LVAD flow scenarios. For all case considered here, when compared against baseline aortic hemodynamics, pulse modulated LVAD flow has a greater extent of vorticity; and constant LVAD flow has lesser extent of vorticity, in the aortic arch. Increased pulsation increased vorticity in the aortic arch, and (except for the *Inc45Azi0* case) in the abdominal aorta as well, although the extent of vorticity in the abdominal aorta is lower than the arch. The extent of vorticity in the aortic arch decreases, and that in the abdominal aorta increases, with increasing angle away from the aortic valve (*Inc* angle). The *Azi* angle also influences the extent of vorticity, in particular, at the aortic arch where the *Azi-45* cases are found to have the highest extent of vorticity for each *Inc* angle.

### 3.3 Wall shear stress variation along the aorta

The computed time averaged wall shear stress (as described in Section 2.3) for all 27 cases of LVAD nasto-mosis considered, are illustrated in panel a. of Figures 7 and 8 for the aortic arch and the abdominal aorta respectively. Row and column arrangement with respect to angles and pulsation is the same as Figure 3. The corresponding baseline scenario is illustrated in Figure 2 panel b. The comparison shows elevated WSS in the LVAD outflow jet impingement region, which is strongly influenced by the LVAD outflow graft anastomosis. As the flow progresses post-impingement to the vessel regions downstream, this influence of anastomosis is observed on variations in WSS values in the descending aorta into the abdominal aorta. These observations are further advanced quantitatively, by comparing ratio of the computed ⟨*τ*_*avg*_⟩ metric (as outlined in Section 2.3) for each of the 27 LVAD flow scenarios with respect to that for the baseline flow. This is illustrated in panel b. in Figures 7 and 8 for the aortic arch wall, and the abdominal aorta wall respectively. Additionally, to further illustrate the jet impingement influence on WSS distribution, the descriptor ⟨Δ*τ*_*avg*_⟩ as defined in Equation 15 was computed, and scaled by the overall maximum TAWSS value (153 dyn/cm^2^) observed for the 28 cases including the baseline case. The resultant surface visualization is presented in Figure 7, panel c., which depicts that the highest normalized differences are located in the region of LVAD outflow jet impingement onto the aorta wall, which is strongly dependent on the surgical anastomosis angles. The regions distal from the impingement zone have small TAWSS differences from baseline. To characterize the impingement neighborhood in greater detail, we also conducted a quantitative comparison of TAWSS difference using the descriptor 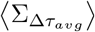 as defined in Equation 16. The resultant values for 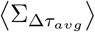 are visualized in Figure 7 panel d. for all 27 LVAD flow scenarios considered. For the aortic arch, for 7 out of the 9 anastomosis considered, constant flow has greater difference in TAWSS from baseline, compared to pulse modulated cases. However, influence of pulse modulation is less pronounced compared to that of anastomosis angles, where both *Inc* and *Azi* angles influence the TAWSS compared to baseline. Specifically, *Azi-45* angles present some of the lowest TAWSS values in the arch compared to baseline, while *Inc135Azi0* anastomosis has the highest TAWSS difference from baseline case. Additionally, when differences are compared pointwise (using 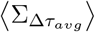), all LVAD cases considered show greater TAWSS in the aortic arch than the baseline, which is expected based on LVAD outflow jet impingement considerations. However, in the abdominal aorta, the influence of pulse modulation gets more pronounced, and TAWSS distribution (compared to baseline flow) increases with increasing pulse modulation. Additionally, TAWSS ratio against baseline flow generally reduces across *Azi0, Azi45*, and *Azi-45* respectively; with all *Azi0* anastomosis showing TAWSS ratio greater than baseline.

**Figure 7:**
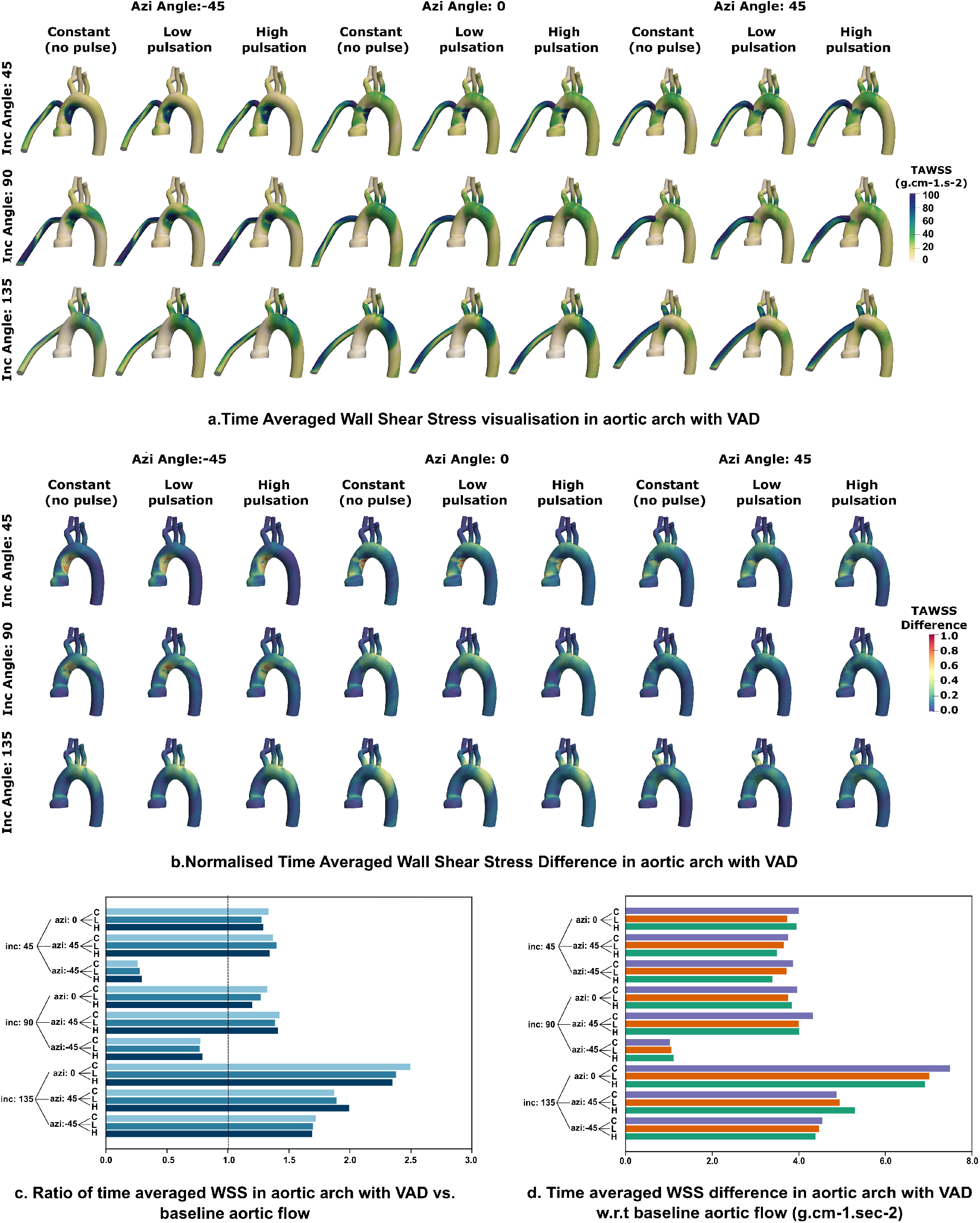
Simulated time averaged wall shear stress (TAWSS) for the aortic arch region for all 27 LVAD flow scenarios. Panel a. presents the actual TAWSS distribution along the aorta wall; while panel c. presents the distribution of the normalized wall shear difference from baseline ⟨Δτ_avg_⟩ (as defined in Section 3.3). Panels b. and d. present the ratio of the surface averaged descriptor ⟨τ_avg_⟩, and the averaged wall shear difference descriptor 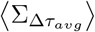. Ratio=1.0 in panel b. represents baseline flow without LVAD.

**Figure 8:**
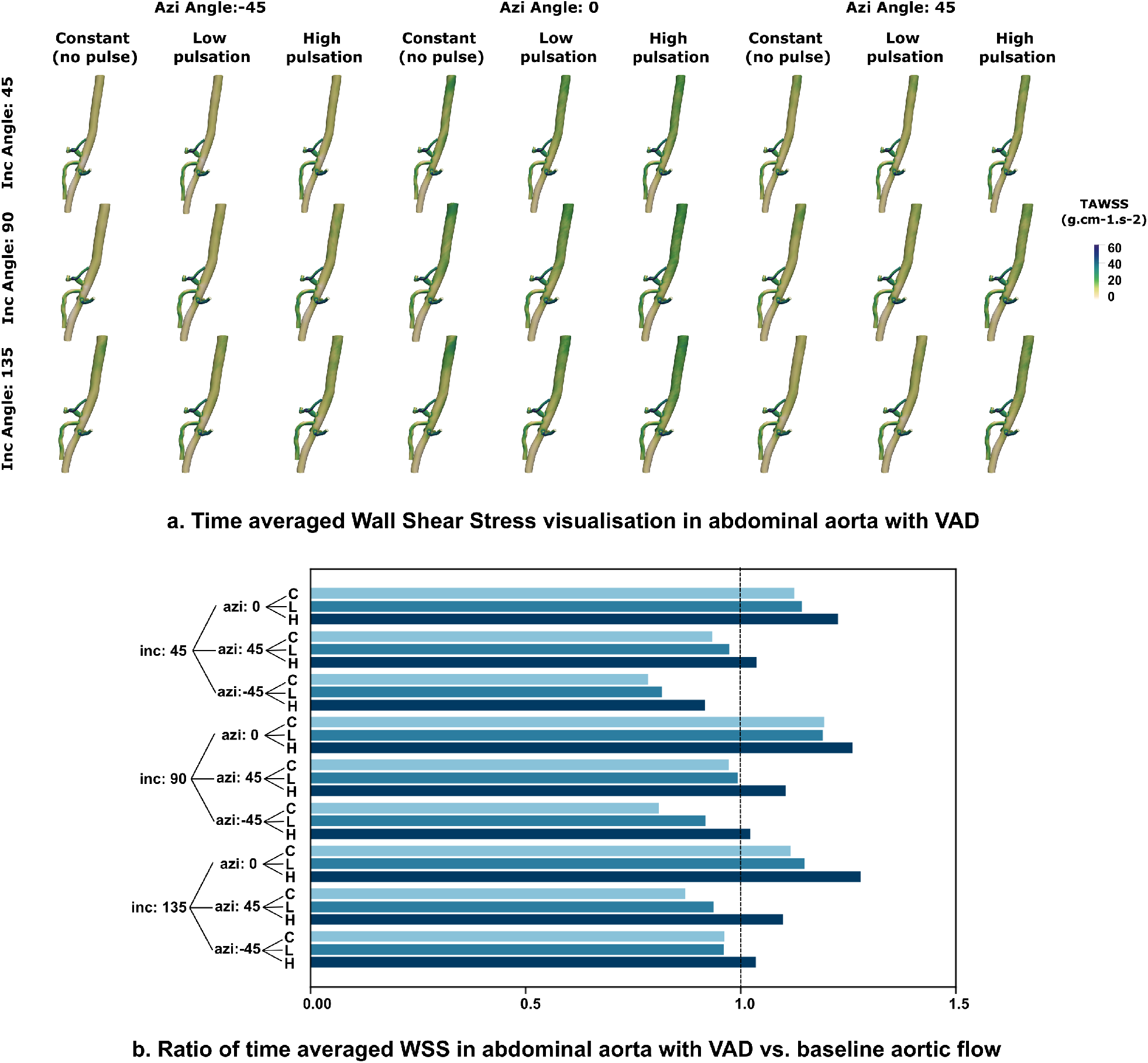
Simulated time averaged wall shear stress (TAWSS) for the abdominal aorta for all 27 LVAD flow scenarios. Panel a. presents the actual TAWSS distribution along the aorta wall; while panel b. present the ratio of the surface averaged descriptor ⟨τ_avg_⟩ compared to baseline flow. Ratio=1.0 in panel b. represents baseline flow without LVAD.

### 3.4 Quantifying the extent of hemodynamic alteration from baseline

For the helicity descriptor ⟨Φ_*H*_⟩, vorticity descriptor ⟨Φ_*Q*_⟩, and wall shear stress descriptor ⟨*τ*_*avg*_⟩, leading to a total of 8 descriptors for aortic arch and abdominal aorta, we present their ratios compared to baseline flow values with respect to each of the three LVAD parameters (*Inc* angle, *Azi* angle, and pulsation) in Figure 9. As described in Sections 3.1–3.3, large deviations of these ratios from unity indicate significant deviations from pre-LVAD baseline flow. We observe that across all three parametric variations, helicity descriptors ⟨Φ_*H*_⟩ for both the arch and abdominal aorta, and wall shear descriptors ⟨*τ*_*avg*_⟩ for abdominal aorta are in general closer to 1.0, while vorticity descriptors ⟨Φ_*Q*_⟩ for both arch and abdominal aorta, and wall shear descriptors for aortic arch show greater departure from unity. When averaged across all 9 surgical anastomosis considered, increased pulsation shows: (a) consistent reduction in helical flow for both arch and abdominal aorta, bringing them closer to baseline flow (that is, ratio = 1.0); (b) consistent increase in abdominal aorta wall shear; and (c) increase in vorticity for both arch and abdominal aorta. Anastomosis angles show a strong influence in terms of changing wall shear in aortic arch, and vorticity in both arch and abdominal aorta to deviate from baseline flow. Conversely, wall shear at abdominal aorta deviates from baseline to a lesser extent with varying anastomosis. We synthesize all of these observations to quantitatively identify which flow scenario shows the greatest/least extent of alteration from baseline. For this, we sum up the deviation from unity for each of these descriptor ratios against baseline, to create a cumulative quantitative metric for the extent of hemodynamic alteration, defined as follows:

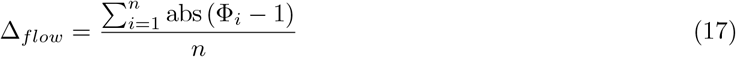

**Figure 9:**
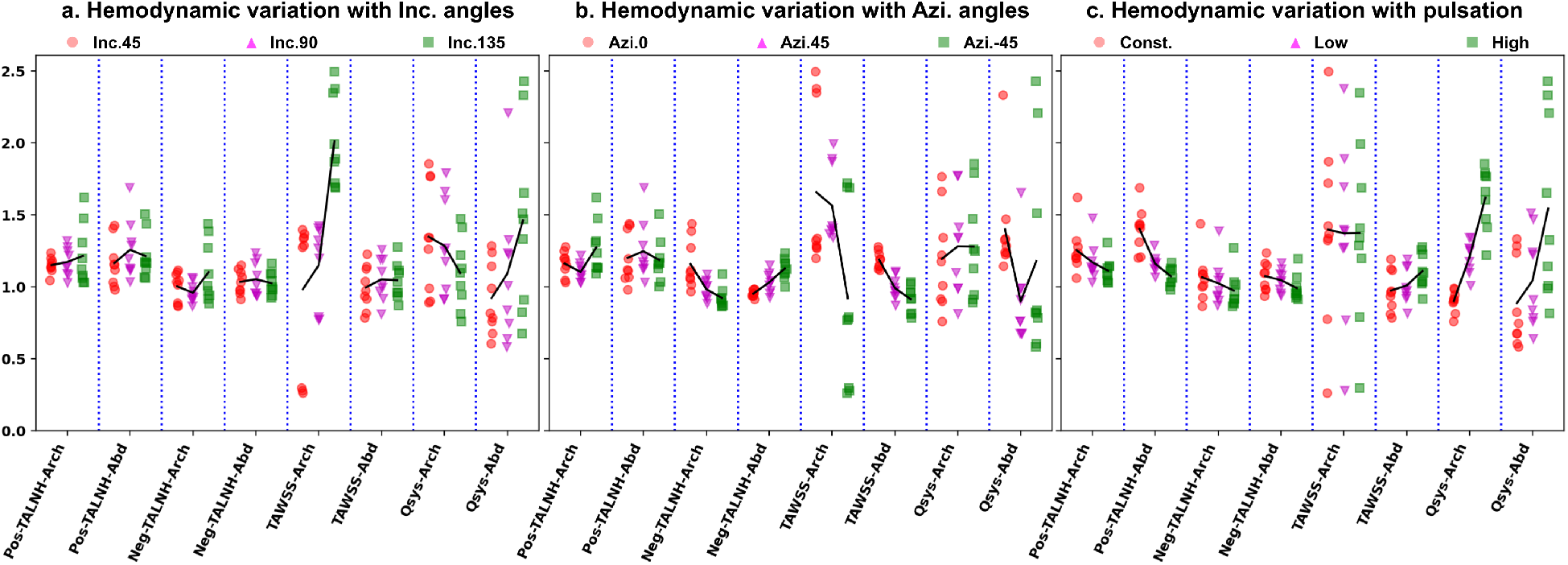
Factorial illustration of computed data for all 8 hemodynamic descriptors described in this study, to illustrate the extent of altered hemodynamics due to LVAD operation, compared to pre-LVAD baseline flow. Panel a. presents variation of the hemodynamic descriptors with the outflow graft angle towards/away from aortic valve (Inc. angle). Panel b. presents the same for varying graft angles to the left/right of the heart (Azi. angles). Panel c. presents the influence of pulse modulation for flow across all graft surgical anastomosis considered.

where Φ_*i*_ is a placeholder for the helicity descriptor ratio, wall shear descriptor ratio, and vorticity descriptor ratio; and *n* denotes the number of such descriptors being computed. Considering all the 8 descriptors as illustrated in Figure 9, the lowest Δ_*flow*_ value is for low pulse modulated flow in *Inc90AziNeg45* anastomosis model, while the highest Δ_*flow*_ value is for high pulse modulated flow in *Inc135Azi0* anastomosis model. We note that while this quantitative analysis does provide insights into the amount of hemodynamic alteration from baseline due to LVAD implantation, this analysis alone will not sufficiently indicate whether certain anastomosis or Δ_*flow*_ level is correlated with a specific pathology.

## 4 Discussion

### 4.1 A two jet flow model for LVAD hemodynamics

We interpret the observations from our analysis using the premise of flow physics originating due to two different jets and their consequent flow patterns. For normal physiology (without VAD-support), aortic hemodynamics is driven by a jet flow emanating from aortic valve opening during ventricular systole. This pulsatile *“Aortic outflow jet”* (AO jet) is directed along the aorta curvature or centerline, and interacts with aorta walls at a location distal from the valve, subsequently determining spatiotemporal flow patterns and state of wall shear. For LVAD operation, aortic hemodynamics is instead driven by the *“LVAD outflow jet”* (LO jet), which traverses across the aorta centerline and impinges directly on the aorta wall. LO Jet impingement and flow roll-up leads to significantly different vortex generation and flow patterns compared to AO jet, the vortices subsequently moving from the impingement site into the descending and abdominal aorta. While LO jet flow occurs throughout the cardiac cycle, physiological AO jet is active during systole only. Under high LVAD support, most commonly the aortic valve is closed, there is no AO jet flow, and only LO jet flow is active ^[27,15]^. This corresponds to the scenario considered in our study. However, in case of intermittent aortic valve reopening under high trans-valvular pressure, a weak form of the AO jet can interplay with the LO jet, contributing further to the spatiotemporal complexity of aortic hemodynamics ^[29]^. Flow due to intermittent aortic valve opening has not been considered here to keep within the scope of the study, but the approach provided here can be directly extended to evaluate how jet interaction effects influence hemodynamic alterations. Nevertheless, based on observations presented here, we can reason that the differences of these two jet flows (and their potential interactions during intermittent valve opening) ultimately determines the altered state of hemodynamics post-LVAD implantation. Consequently, any variable affecting the two jets can play a potential role in influencing flow and flow-mediated pathologies in patients. Together this framework leads to the important distinction of whether: (a) the LVAD flow itself, or (b) the extent by which LVAD flow differs from baseline; is more relevant to understand LVAD treatment outcomes and etiology of post-treatment complications. This requires and motivates further investigation in future studies.

We can further compile three key flow physics interpretations in accordance with this two jet model. First, while LO jet impingement primarily drives the aortic flow, the aortic geometry, curvature, and tortuosity, also play a concurrent role in ultimately determining the altered state of hemodynamics. Specifically, curvature induced secondary vortices are known to shape helical flow patterns in the aorta. Impingement induced flow is likely to be more positive helical, while baseline flow tendency is more negative helical. The actual extent of helicity and mixing in a patient with LVAD will likely be a balance of the two. For scenarios where aortic geometry effects gain prominence, negative helicity increases, thereby explaining some of the trends in helicity variation with anastomosis as observed from our simulations. One parameter that can enable further characterization of such effects is the Dean number calculated based on aortic curvature. For all LVAD models considered, the Dean number computed based on maximum flow velocity in the arch ranged between 850-2232, whereas the same for the baseline flow was 9346. Especially considering that real aortic arch is not a simple curved tube, but has added geometric complexities, the relative magnitudes of arch-based Dean number can further identify the strength of secondary circulatory flow originating from LO jet impingement. Second, flow development post-impingement in the aortic arch determines altered state of hemodynamics elsewhere. For example, LO jet impingement will cause significantly greater extent of vorticity in the arch compared to baseline. However, as flow traverses distally into the descending aorta, this vorticity dissipates continuously due to viscosity. Thus, at shallower *Inc* angles, vortices due to LO jet impingement originate closer to aortic valve, and by the time flow reaches abdominal aorta, they undergo a higher amount of dissipation, reducing their influence on the flow. Third, the LO jet impingement hemodynamics is influenced by both anastomosis angle and pulsation, through different modalities. The former influences the geometry or angle of impingement, while the latter influences the temporal intensity of the impingement flow. This is evident from comparing factors such as vorticity and state of wall shear locally at the arch and distally at the abdominal aorta.

### 4.2 Study implications and clinical relevance

Here we have characterized the hemodynamics due to LVAD flow compared to baseline flow, as function of outflow graft anastomosis and pulsation. While the anastomosis angle towards/away from valve (*Inc* angle) has been extensively studied ^[10,4]^, here we provide insights on how the angle to left/right of heart (*Azi* angle) can also play a role that is less explored. The resulting altered flow patterns in the arch originating due to LVAD jet impingement can determine transport and fate of thromboemboli entering the arch from the graft, causing subsequent ischemic stroke ^[41,43]^. Flow stasis in the aortic root, as described here, can also lead to thrombogenicity, and further increase the risk of ischemic stroke or other embolic events in downstream circulation ^[4]^. Flow vorticity and helicity are also widely acknowledged to play a role in promoting mixing in aortic hemodynamics ^[36]^. The state of shear stress is critical as well, due to the increasingly acknowledged role of overall shear forces in elevating the risk of GI bleeding in LVAD patients ^[40,7]^. This study is one of the first attempts to analyze the influence of LVAD outflow jet impingement for varying surgical angles and pulsation, at vascular locations distal from the graft entry. Specifically, hemodynamics in the abdominal aorta is acknowledged to play a key role in determining risk for GI bleeding, yet has been less commonly studied compared to the aortic arch. We observe that all three variables considered here influenced wall shear at abdominal aorta. E.g. abdominal aorta TAWSS increased with increasing pulsation, and reduced with *Azi* angles 0, 45°, and −45° respectively. Additional specifics regarding the role of shear stresses on von Willebrand Factor (vWF), circulating vWF degradation, and hemodynamic connection to vWF disease in bleeding requires further investigations. There is also evidence of wall shear being of influence in hormonal regulation and endocrine signaling which can be further explored. Furthermore, we discussed in detail the role of introducing pulse modulation in continuous LVAD flows. Pulse modulation was shown to promote increased vorticity, and reduce flow stasis, which can further influence risk of thromboembolic events. This provides additional insight into benefits of pulse-modulation in LVAD patients as outlined in other works ^[44]^. Thus, the framework devised here lays the basis for future discussion on clinical correlation of LVAD graft angles and pulsation with actual stroke and GI bleeding risk quantifiers; which is of high clinical relevance.

### 4.3 Assumptions and limitations

This *in silico* study was based on several key assumptions, with associated limitations, as discussed here. First, we note that hemodynamic descriptors such as the ones used here, are a single unified quantifier that can differentiate the spatiotemporal complexity of the bulk flow characteristics for different LVAD flow cases. They are not suitable for comparing point-wise flow variables for varying LVAD parameters, neither was that the goal. Instead, the objective is to develop a reduced order space of quantifiers from the 3D space-time varying flow data, to compare the effect of parametric variations in surgical anastomosis and flow modulation for LVAD flows. To this end, helicity, vorticity, and wall shear, are fundamental hemodynamic descriptors used in this study; while there are a wide range of additional descriptors such as oscillatory shear descriptors, transverse shear descriptors, Lagrangian residence time, Finite-time Lyapunov Exponents (FTLE), etc. which can be incorporated to further characterize LVAD hemodynamics. Second, we assumed here that the aortic valve was always shut. However, intermittent opening of aortic valve is observed in LVAD patients, and can significantly alter the state of hemodynamics as per the two-jet interpretation in Section 4.1; which was not accounted for this study. Similarly, this was a purely hemodynamics based LVAD study and the arterial wall response to the LVAD induced hemodynamics needs further investigation. Each of these aspects can involve sufficient computational modeling complexity to necessitate a separate study itself. Third, specifically in terms of hemodynamics, complexities such as non-Newtonian hemorheology, viscoelastic effects, and flow transition into turbulence locally in the jet impingement zone may necessitate improved higher-order hemodynamic modeling to accurately describe higher resolution flow patterns and wall shear stresses due to LVAD jet impingement. Fourth, the base vasculature model was from a single healthy subject, on which LVAD grafts were virtually attached. A comprehensive analysis considering multiple subjects to account for patient-to-patient variations was not within the scope of this study. Lastly, additional LVAD related parameters such as ventricular coupling or surgical location along the aorta, have not been considered in detail, also primarily to keep within the scope of the study.

### 4.4 Concluding remarks

We have developed a quantitative framework for hemodynamic assessment in patient-specific vasculature with an operating LVAD. Specifically, the framework comprised: (a) obtaining an estimate of what aortic hemodynamics would be without LVAD support (*baseline flow*); (b) obtaining an estimate of what aortic hemodynamics is with LVAD support; and (c) characterizing the LVAD operation in terms of differences between hemodynamics computed in (a), and (b). This is referred to as hemodynamic alterations from baseline flow. We have demonstrated this framework by conducting a parametric study to quantify how LVAD outflow graft angles and pulse modulation influences hemodynamic alterations as defined above. For this, we have specifically quantified alterations in terms of three key flow features: helical flow, vorticity, and wall shear stress. Results from this parametric study spanning 28 different CFD models clearly establishes that our framework can: (a) identify how specific LVAD parameters (such as graft angle, and pulsation) can influence the altered state of aortic hemodynamics compared to baseline; and (b) identify combinations of LVAD parameters with the highest or lowest amount of hemodynamic alterations. Lastly, our observations were compiled into a conceptual two jet model for generalized assessment of aortic hemodynamics in a patient with an operating LVAD.

## Supporting information

Supplemental animation: Q-combined-anim.mp4

Supplemental animation: vel-combined-anim.mp4

## Data Availability

All data produced in the present work are contained in the manuscript. Human vascular model data obtained from open source Vascular Model repository (www.vascularmodel.com) associated with the open source SimVascular project (simvascular.github.io)

## 6 Acknowledgements

This work was supported by a University of Colorado Anschutz-Boulder (AB) Nexus Research Collaboration Grant awarded to authors DM and JP. This work utilized resources from the University of Colorado Boulder Research Computing Group, which is supported by the National Science Foundation (awards ACI-1532235 and ACI-1532236), the University of Colorado Boulder, and Colorado State University. AS conducted the modeling, simulation, and quantitative analysis for the study. DM designed the study, conducted data analysis and interpretation, and drafted the manuscript in collaboration with AS, EM, and JP. JP and EM guided the study design, and hemodynamic parameter selection alongwith clinical interpretation of the data. All four authors reviewed and finalized the draft, and are in agreement regarding the final content of the manuscript.

## Supplementary Material Information

### Animation of hemodynamic patterns

Along with the manuscript we have included two separate animation files. The first animation file included with the name *“vel-combined-anim*.*mp4”* presents a combined animation of blood flow velocity in the aortic arch for all 27 LVAD flow scenarios considered in this study. The second animation file included with the name *“Q-combined-anim*.*mp4”* presents likewise an animation of vortical structures identified by Q criterion isosurfaces (as described in the manuscript) for all 27 LVAD flow scenarios considered. Both of these animations provide a clear view of the dynamics of the LVAD outflow jet impingement onto the aorta wall. Each animation is 10 seconds long, and considering the pulse cycle as 1 second, each animation is therefore rendered 10 times slower than actual timescales.

### Comparing hemodynamic descriptors between arch and abdominal aorta

Here, we present an additional analysis based on computed hemodynamic descriptor data, to illustrate how the altered state of flow due to LVAD operation compares between the aortic arch and the abdominal aorta region. For this, we compute the ratio of the helicity descriptor ⟨Φ_*H*_⟩, and the time averaged wall shear stress descriptor ⟨*τ*_*avg*_ ⟩, between the aortic arch and the abdominal aorta; and compare them against the baseline. These are illustrated in Figure S1, where panel a. depicts the ratios for LNH at +0.2; panel b. presents the ratios for LNH at −0.2; and panel c. presents the ratios for the wall shear descriptor. To enable comparing with the baseline, the corresponding hemodynamic descriptor ratios are also computed for the baseline flow. It is observed that for all of the 27 LVAD flow scenarios considered, the ratios are greater than the corresponding baseline values. Specifically, extent of negative helicity is more commonly greater in the aortic arch than the abdominal aorta, while extent of positive helicity is more commonly lesser in the aortic arch than the abdominal aorta. For all the *Inc135* angle cases, the wall shear in the aortic arch is greater than the abdominal aorta. This follows the explanation provided in the main manuscript, where for increasing the angle away from the aortic root delays impingement and causes the LVAD outflow jet to roll into the descending aorta into the abdominal aorta.

### Visualizing raw data for hemodynamic alteration

Here, we present an additional visualization of the computed raw data for the cumulative metric for hemodynamic alteration as outlined in Equation 17 in Section 3.4. These have been presented in Figure S2. Specifically, the computed numerical values for the parameter Δ_*flow*_ for each of the 27 LVAD flow scenarios considered have been tabulated, and then colored on a gradient scale in green based on the values. Darker green would indicate high Δ_*flow*_ values. It can be seen directly based on Figure S2 that the cases with the highest and lowest values for Δ_*flow*_ are the ones reported in Section 3.4.

**Figure S1:**
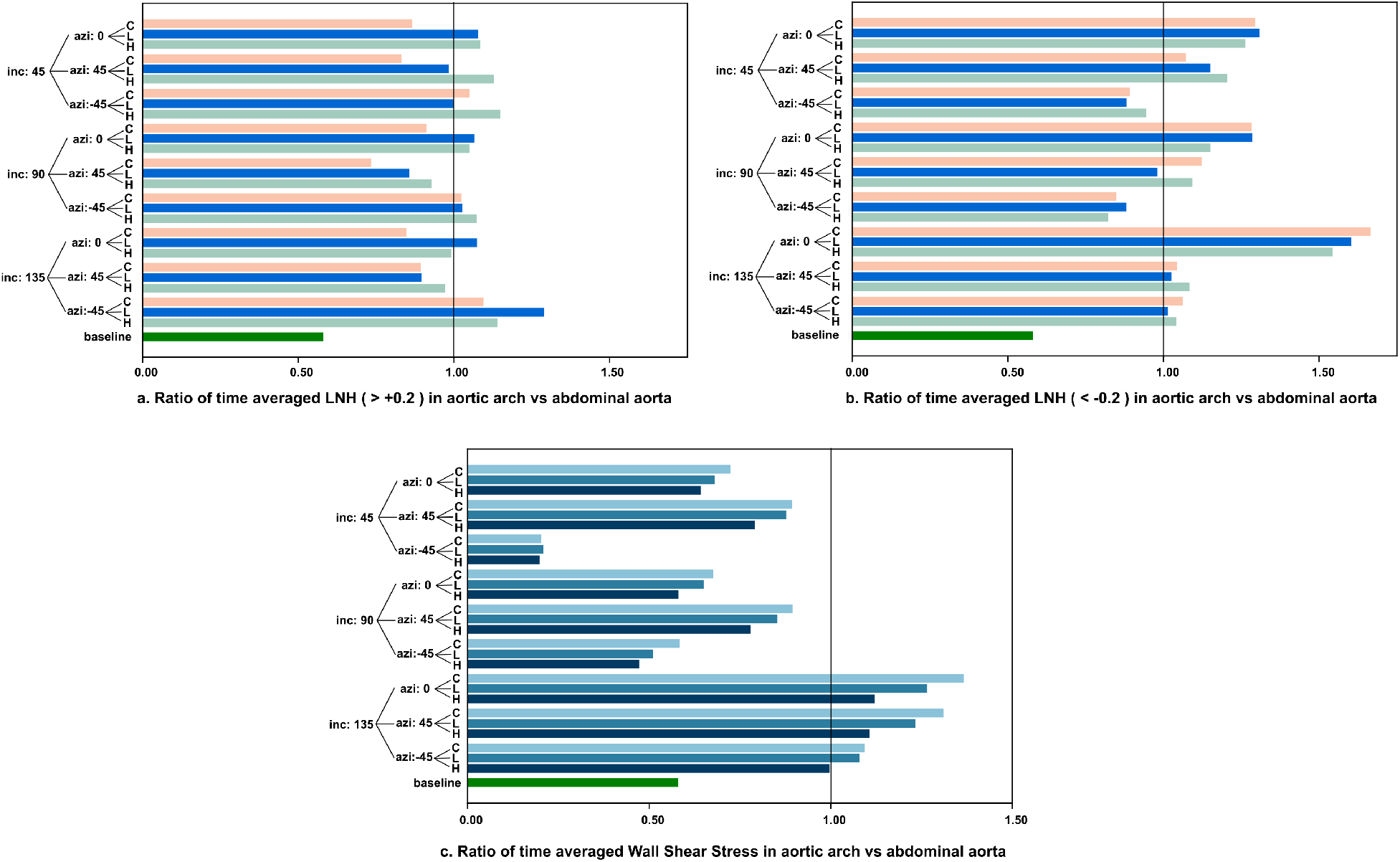
Ratios of computed flow descriptors for aortic arch vs abdominal aorta. Panel a. presents ratios for positive helicity descriptor; panel b. presents ratios for negative helicity descriptor; and panel c. presents ratios for wall shear stress descriptor. Corresponding values for baseline are included in each panel for comparison.

**Figure S2:**
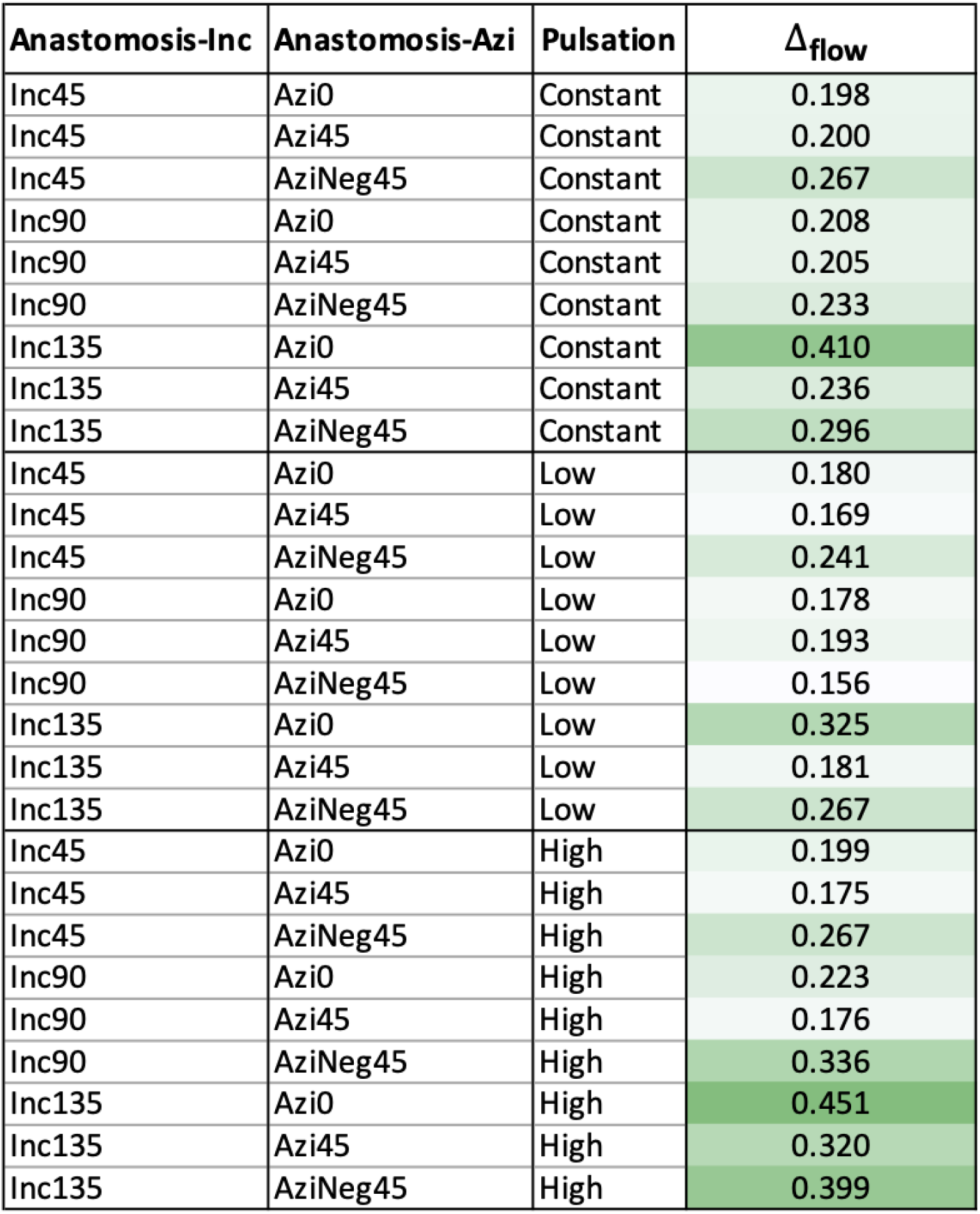
Panel presenting computed Δ_flow_ values for all 27 LVAD flow scenarios considered.

## References

[1] D. Acharya, R. Loyaga-Rendon, C. J. Morgan, K. A. Sands, S. V. Pamboukian, I. Rajapreyar, W. L. Holman, J. K. Kirklin, and J. A. Tallaj. Intermacs analysis of stroke during support with continuous-flow left ventricular assist devices: risk factors and outcomes. JACC: Heart Failure, 5(10):703–711, 2017.

[2] J. Ahrens, B. Geveci, and C. Law. Paraview: An end-user tool for large-data visualization. In C. D. Hansen and C. R. Johnson, editors, Visualization Handbook, pages 717–731. Butterworth-Heinemann, Burlington, 2005.

[3] J. Alastruey, J. H. Siggers, V. Peiffer, D. J. Doorly, and S. J. Sherwin. Reducing the data: Analysis of the role of vascular geometry on blood flow patterns in curved vessels. Physics of Fluids, 24(3):031902, 2012.

[4] A. Aliseda, V. K. Chivukula, P. Mcgah, A. R. Prisco, J. A. Beckman, G. J. Garcia, N. A. Mokadam, and C. Mahr. Lvad outflow graft angle and thrombosis risk. ASAIO Journal (American Society for Artificial Internal Organs: 1992), 63(1):14, 2017.

[5] A. Arzani and S. C. Shadden. Characterizations and correlations of wall shear stress in aneurysmal flow. Journal of Biomechanical Engineering, 138(1):014503, 2016.

[6] C. R. Bartoli, G. A. Giridharan, K. N. Litwak, M. Sobieski, S. D. Prabhu, M. S. Slaughter, and S. C. Koenig. Hemodynamic responses to continuous versus pulsatile mechanical unloading of the failing left ventricle. ASAIO Journal (American Society for Artificial Internal Organs: 1992), 56(5):410–416, 2010.

[7] C. R. Bartoli, D. M. Zhang, S. Hennessy-Strahs, J. Kang, D. J. Restle, C. Bermudez, P. Atluri, and M. A. Acker. Clinical and in vitro evidence that left ventricular assist device–induced von willebrand factor degradation alters angiogenesis. Circulation: Heart Failure, 11(9):e004638, 2018.

[8] E. Benjamin, P. Muntner, A. Alonso, M. Bittencourt, C. Callaway, A. Carson, A. Chamberlain, A. Chang, S. Cheng, S. Das, F. Delling, et al. Heart Disease and Stroke Statistics – 2019 Update. Circulation, 139, 2019.

[9] E. Braunwald. Cardiovascular medicine at the turn of the millennium: triumphs, concerns, and opportunities. New England Journal of Medicine, 337(19):1360–1369, 1997.

[10] A. Callington, Q. Long, P. Mohite, A. Simon, and T. K. Mittal. Computational fluid dynamic study of hemodynamic effects on aortic root blood flow of systematically varied left ventricular assist device graft anastomosis design. The Journal of Thoracic and Cardiovascular Surgery, 150(3):696–704, 2015.

[11] M. V. Caruso, V. Gramigna, M. Rossi, G. F. Serraino, A. Renzulli, and G. Fragomeni. A computational fluid dynamics comparison between different outflow graft anastomosis locations of left ventricular assist device (lvad) in a patient-specific aortic model. International Journal for Numerical Methods in Biomedical Engineering, 31(2):e02700, 2015.

[12] Centers for Disease Control and Prevention, National Center for Health Statistics. Underlying cause of death, 1999–2017. https://wonder.cdc.gov/ucd-icd10.html, 2019.

[13] Z. Chen, S. K. Jena, G. A. Giridharan, M. A. Sobieski, S. C. Koenig, M. S. Slaughter, B. P. Griffith, and Z. J. Wu. Shear stress and blood trauma under constant and pulse-modulated speed cf-vad operations: Cfd analysis of the hvad. Medical & Biological Engineering & Computing, 57(4):807–818, 2019.

[14] G. Coppola and C. Caro. Oxygen mass transfer in a model three-dimensional artery. Journal of The Royal Society Interface, 5(26):1067–1075, 2008.

[15] J. G. da Rocha e Silva, A. L. Meyer, S. Eifert, J. Garbade, F. W. Mohr, and M. Strueber. Influence of aortic valve opening in patients with aortic insufficiency after left ventricular assist device implantation. European Journal of Cardio-Thoracic Surgery, 49(3):784–787, 2016.

[16] K. H. Fraser, M. E. Taskin, B. P. Griffith, and Z. J. Wu. The use of computational fluid dynamics in the development of ventricular assist devices. Medical Engineering & Physics, 33(3):263–280, 2011.

[17] L. J. Frazin, M. J. Vonesh, K. B. Chandran, T. Shipkowitz, A. S. Yaacoub, and D. D. McPherson. Confirmation and initial documentation of thoracic and abdominal aortic helical flow. an ultrasound study. ASAIO Journal (American Society for Artificial Internal Organs: 1992), 42(6):951–956, 1996.

[18] D. Gallo, D. A. Steinman, P. B. Bijari, and U. Morbiducci. Helical flow in carotid bifurcation as surrogate marker of exposure to disturbed shear. Journal of Biomechanics, 45(14):2398–2404, 2012.

[19] J. Garcia, A. J. Barker, J. D. Collins, J. C. Carr, and M. Markl. Volumetric quantification of absolute local normalized helicity in patients with bicuspid aortic valve and aortic dilatation. Magnetic Resonance in Medicine, 78(2):689–701, 2017.

[20] B. C. Good, S. Deutsch, and K. B. Manning. Continuous and pulsatile pediatric ventricular assist device hemodynamics with a viscoelastic blood model. Cardiovascular engineering and technology, 7(1):23–43, 2016.

[21] L. Harvey, C. Holley, S. S. Roy, P. Eckman, R. Cogswell, K. Liao, and R. John. Stroke after left ventricular assist device implantation: outcomes in the continuous-flow era. The Annals of Thoracic Surgery, 100(2):535–541, 2015.

[22] T. Hasin, Y. Matsuzawa, R. R. Guddeti, T. Aoki, T.-G. Kwon, S. Schettle, R. J. Lennon, R. G. Chokka, A. Lerman, and S. S. Kushwaha. Attenuation in peripheral endothelial function after continuous flow left ventricular assist device therapy is associated with cardiovascular adverse events. Circulation Journal, 79(4):770–777, 2015.

[23] G. Inci and E. Sorgüven. Effect of lvad outlet graft anastomosis angle on the aortic valve, wall, and flow. ASAIO Journal (American Society for Artificial Internal Organs: 1992), 58(4):373–381, 2012.

[24] M. Ising, S. Warren, M. A. Sobieski, M. S. Slaughter, S. C. Koenig, and G. A. Giridharan. Flow modulation algorithms for continuous flow left ventricular assist devices to increase vascular pulsatility: a computer simulation study. Cardiovascular Engineering and Technology, 2(2):90, 2011.

[25] H. R. Jabbar, A. Abbas, M. Ahmed, C. T. Klodell, M. Chang, Y. Dai, and P. V. Draganov. The incidence, predictors and outcomes of gastrointestinal bleeding in patients with left ventricular assist device (lvad). Digestive Diseases and Sciences, 60(12):3697–3706, 2015.

[26] J. Jeong and F. Hussain. On the identification of a vortex. Journal of Fluid Mechanics, 285:69–94, 1995.

[27] R. John, K. Mantz, P. Eckman, A. Rose, and K. May-Newman. Aortic valve pathophysiology during left ventricular assist device support. The Journal of Heart and Lung Transplantation, 29(12):1321–1329, 2010.

[28] C. Karmonik, S. Partovi, M. Loebe, B. Schmack, A. Weymann, A. B. Lumsden, M. Karck, and A. Ruhparwar. Computational fluid dynamics in patients with continuous-flow left ventricular assist device support show hemodynamic alterations in the ascending aorta. The Journal of Thoracic and Cardiovascular Surgery, 147(4):1326–1333, 2014.

[29] C. Mahr, V. Chivukula, P. McGah, A. R. Prisco, J. A. Beckman, N. A. Mokadam, and A. Aliseda. Intermittent aortic valve opening and risk of thrombosis in vad patients. ASAIO Journal (American Society for Artificial Internal Organs: 1992), 63(4):425, 2017.

[30] D. Mancini and P. C. Colombo. Left ventricular assist devices: a rapidly evolving alternative to transplant. Journal of the American College of Cardiology, 65(23):2542–2555, 2015.

[31] J. Marsano, J. Desai, S. Chang, M. Chau, M. Pochapin, and G. E. Gurvits. Characteristics of gastrointestinal bleeding after placement of continuous-flow left ventricular assist device: a case series. Digestive Diseases and Sciences, 60(6):1859–1867, 2015.

[32] K. May-Newman, B. Hillen, C. Sironda, and W. Dembitsky. Effect of lvad outflow conduit insertion angle on flow through the native aorta. Journal of Medical Engineering & Technology, 28(3):105–109, 2004.

[33] A. Millon, M. Sigovan, L. Boussel, J.-L. Mathevet, V. Louzier, C. Paquet, A. Geloen, N. Provost, Z. Majd, D. Patsouris, et al. Low wss induces intimal thickening, while large wss variation and inflammation induce medial thinning, in an animal model of atherosclerosis. PLoS One, 10(11):e0141880, 2015.

[34] T. L. Molina, J. C. Krisl, K. R. Donahue, and S. Varnado. Gastrointestinal bleeding in left ventricular assist device: octreotide and other treatment modalities. ASAIO Journal (American Society for Artificial Internal Organs: 1992), 64(4):433–439, 2018.

[35] U. Morbiducci, R. Ponzini, M. Grigioni, and A. Redaelli. Helical flow as fluid dynamic signature for atherogenesis risk in aortocoronary bypass. a numeric study. Journal of Biomechanics, 40(3):519–534, 2007.

[36] U. Morbiducci, R. Ponzini, G. Rizzo, M. Cadioli, A. Esposito, F. M. Montevecchi, and A. Redaelli. Mechanistic insight into the physiological relevance of helical blood flow in the human aorta: an in vivo study. Biomechanics and Modeling in Mechanobiology, 10(3):339–355, 2011.

[37] J. A. Morgan, R. J. Brewer, H. W. Nemeh, B. Gerlach, D. E. Lanfear, C. T. Williams, and G. Paone. Stroke while on long-term left ventricular assist device support: incidence, outcome, and predictors. ASAIO Journal (American Society for Artificial Internal Organs: 1992), 60(3):284–289, 2014.

[38] D. Mukherjee and S. C. Shadden. Inertial particle dynamics in large artery flows–implications for modeling arterial embolisms. Journal of Biomechanics, 52:155–164, 2017.

[39] D. Mukherjee, J. Padilla, and S. C. Shadden. Numerical investigation of fluid–particle interactions for embolic stroke. Theoretical and Computational Fluid Dynamics, 30(1-2):23–39, 2016.

[40] A. Nascimbene, S. Neelamegham, O. Frazier, J. L. Moake, and J.-f. Dong. Acquired von willebrand syndrome associated with left ventricular assist device. Blood, The Journal of the American Society of Hematology, 127(25):3133–3141, 2016.

[41] A. F. Osorio, R. Osorio, A. Ceballos, R. Tran, W. Clark, E. A. Divo, I. R. Argueta-Morales, A. J. Kassab, and W. M. DeCampli. Computational fluid dynamics analysis of surgical adjustment of left ventricular assist device implantation to minimise stroke risk. Computer Methods in Biomechanics and Biomedical Engineering, 16(6):622–638, 2013.

[42] N. S. Parikh, J. Cool, M. G. Karas, A. K. Boehme, and H. Kamel. Stroke risk and mortality in patients with ventricular assist devices. Stroke, 47(11):2702–2706, 2016.

[43] R. Prather, E. Divo, A. Kassab, and W. DeCampli. Computational fluid dynamics study of cerebral thromboembolism risk in ventricular assist device patients: Effects of pulsatility and thrombus origin. Journal of Biomechanical Engineering, 143(9), 2021.

[44] S. N. Purohit, W. K. Cornwell III, J. D. Pal, J. Lindenfeld, and A. V. Ambardekar. Living without a pulse: the vascular implications of continuous-flow left ventricular assist devices. Circulation: Heart Failure, 11(6):e004670, 2018.

[45] O. M. Repository. https://www.vascularmodel.com, 2020.

[46] V. L. Roger. Epidemiology of heart failure: a contemporary perspective. Circulation research, 128(10): 1421–1434, 2021.

[47] G. Savarese and L. H. Lund. Global public health burden of heart failure. Cardiac failure review, 3(1): 7, 2017.

[48] P. Shah, M. Yuzefpolskaya, G. W. Hickey, K. Breathett, O. Wever-Pinzon, V. Khue-Ton, W. Hiesinger, D. Koehl, J. K. Kirklin, R. S. Cantor, et al. Twelfth interagency registry for mechanically assisted circulatory support report: readmissions after left ventricular assist device. The Annals of Thoracic Surgery, 113(3):722–737, 2022.

[49] Simvascular. https://simvascular.github.io, 2022.

[50] K. G. Soucy, S. C. Koenig, G. A. Giridharan, M. A. Sobieski, and M. S. Slaughter. Defining pulsatility during continuous-flow ventricular assist device support. The Journal of Heart and Lung Transplantation, 32(6):581–587, 2013.

[51] L. W. Stevenson and E. A. Rose. Left ventricular assist devices: bridges to transplantation, recovery, and destination for whom? Circulation, 108(25):3059–3063, 2003.

[52] S. Tolpen, J. Janmaat, C. Reider, F. Kallel, D. Farrar, and K. May-Newman. Programmed speed reduction enables aortic valve opening and increased pulsatility in the lvad-assisted heart. Asaio Journal, 61(5):540–547, 2015.

[53] A. Updegrove, N. M. Wilson, J. Merkow, H. Lan, A. L. Marsden, and S. C. Shadden. Simvascular: an open source pipeline for cardiovascular simulation. Annals of biomedical engineering, 45(3):525–541, 2017.

[54] J. S. Uzarski, E. W. Scott, and P. S. McFetridge. Adaptation of endothelial cells to physiologically-modeled, variable shear stress. PloS One, 8(2):e57004, 2013.

[55] N. Yang, S. Deutsch, E. G. Paterson, and K. B. Manning. Numerical study of blood flow at the end-to-side anastomosis of a left ventricular assist device for adult patients. Journal of Biomechanical Engineering, 131:111005:1–9, 2009.

[56] N. Yang, S. Deutsch, E. G. Paterson, and K. B. Manning. Comparative study of continuous and pulsatile left ventricular assist devices on hemodynamics of a pediatric end-to-side anastomotic graft. Cardiovascular engineering and technology, 1(1):88–103, 2010.

[57] Q. Zhang, B. Gao, and Y. Chang. Helical flow component of left ventricular assist devices (lvads) outflow improves aortic hemodynamic states. Medical science monitor: international medical journal of experimental and clinical research, 24:869, 2018.

